# Infrequent intranasal oxytocin followed by positive social interaction improves symptoms in autistic children: a pilot randomized clinical trial

**DOI:** 10.1101/2022.01.03.22268708

**Authors:** Jiao Le, Lan Zhang, Weihua Zhao, Siyu Zhu, Chunmei Lan, Juan Kou, Qianqian Zhang, Yingying Zhang, Qin Li, Zhuo Chen, Meina Fu, Christian Montag, Rong Zhang, Wenxu Yang, Benjamin Becker, Keith M. Kendrick

## Abstract

There are currently no approved drug interventions for social behavior dysfunction in autism spectrum disorder (ASD). Previous trials investigating effects of daily intranasal oxytocin treatment have reported inconsistent results and have not combined it with positive social interaction. However, In two preclinical studies we established that treatment every-other-day rather than daily is more efficacious in maintaining neural and behavioral effects by reducing receptor desensitization. We aimed to establish whether a 6-week intranasal oxytocin compared with placebo treatment, followed by a period of positive social interaction, would produce reliable symptom improvements in children with ASD. A pilot double-blind, randomized, crossover design trial was completed including 41 children with ASD aged 3-8 years. Primary outcomes were the Autism Diagnostic Observation Schedule-2 (ADOS-2) and social responsivity scale-2 (SRS-2). Secondary measures included cognitive, autism and caregiver-related questionnaires and social attention assessed using eye-tracking. Significant improvements were found for oxytocin relative to placebo in primary outcome measures (total ADOS-2 and SRS-2 scores, ps < 0.001) and in behavioral adaptability and repetitive behavior secondary measures. Altered SRS-2 scores were associated with increased saliva oxytocin concentrations. Additionally, oxytocin significantly increased time spent viewing dynamic social compared to geometric stimuli and the eyes of angry, happy and neutral expression faces. There were no adverse side-effects of oxytocin treatment. Overall, results demonstrate that a 6-week intranasal oxytocin treatment administered every other day and followed by positive social interactions can improve clinical, eye-tracking and questionnaire-based assessments of symptoms in young autistic children.

## Introduction

Autism spectrum disorder (ASD) is a neurodevelopmental disorder with core symptoms including social communication and interaction problems and restricted and repetitive behavior patterns [1] and a worldwide prevalence around 1% [2,3]. Currently there is no frontline medication for its core social symptoms. A promising therapeutic target to emerge is the hypothalamic neuropeptide oxytocin (OXT). Single doses of intranasal OXT can enhance social cognition and motivation and promote extensive neural effects in both healthy and ASD adults [4-8], and increase visual attention towards social stimuli, the eyes of emotional faces [9-11] and gaze-following [12] all of which are impaired in ASD [13,14]. Peripheral OXT concentrations are lower in children with ASD [15], and OXT receptor (OXTR) polymorphisms are associated with ASD and autistic traits [16,17]. However, clinical trials using repeated daily doses of intranasal OXT for up to 24 weeks have reported modest improvements in social responsivity [18-22] or repetitive/adaptive behaviors [23-25] or no effect [26-30], although with no serious adverse effects.

Previous trials using chronic intranasal OXT administration have adopted a once- or twice-daily dosing frequency with the objective of raising basal concentrations. However, there is no empirical support for this treatment strategy and G protein-coupled receptors exhibit rapid desensitization and down-regulation following exogenous treatment with agonist ligands [32]. Chronic/daily OXT treatment leads to extensive OXTR and vasopressin-1A receptor down-regulation in the rat forebrain [33-36]. Additionally, when OXT binds to its receptor it can differentially recruit intracellular G protein-coupled pathways with increasing OXT bioavailability shifting coupling away from the excitatory Gq-protein to the inhibitory Gi-one [37-39]. This may explain inverted U-shaped dose-response curves reported for acute OXT treatment with higher doses having opposite effects to lower ones [40,41]. Chronic relative to acute dosing with OXT can impair rather than enhance social behavior [42-44] and increase rather than decrease anxiety in rodents [45]. In humans, we have shown that once daily treatment with intranasal OXT for 5 days reduces its effects on neural and anxiolytic responses to emotional stimuli, whereas they are maintained or enhanced by less frequent administration every other day [46,47]. Thus, OXT given at too great a frequency and/or too high a concentration can reduce or even reverse its functional effects.

Simply increasing lower endogenous OXT concentrations [15] by exogenous treatment alone in autistic individuals may not be the optimal strategy for promoting improved social behavior. Acute administration of intranasal OXT primarily influences task-dependent social attention and motivation and facilitation of learning with positive social feedback [48-50]. Indeed, endogenous OXT treatment may enhance the salience of social stimuli in negative as well as positive contexts which could lead to impaired as opposed to improved social symptoms [51, 52]. Thus, ensuring that OXT is only administered prior to a positive social experience may be optimal [see 52].

In the current trial we therefore administered a standard 24IU dose of intranasal OXT every other day for 6 weeks in young children (3 - 8 years old) with ASD in a randomized placebo (PLC) controlled, crossover design trial where subjects received both OXT and PLC treatments. Additionally, caregivers engaged in positive social interactions with their children after each OXT or PLC dose. Our two primary outcome measures were a gold-standard objective clinical measure (Autistic Diagnostic Observation Schedule – 2; ADOS-2) [53] and the caregiver-based social responsivity scale-2 (SRS-2 total score) [54,55]. A number of secondary outcome measures were also taken including two eye-tracking paradigms as measures of social attention and sensitive to autistic symptoms [13,14,56-58]. Associations between symptom improvements and increased peripheral concentrations of OXT, autism social subtype [59] and OXT receptor genotype [16,17] were also investigated. A range of safety measures were included to monitor any adverse effects.

We hypothesized that the novel infrequent dose regime of intranasal OXT administration in conjunction with positive social interactions would produce reliable improvements in both objective and subjective outcome measures of autistic symptoms.

## Materials and Methods

### Trial design and participants

A total of 46 eligible autistic children were enrolled in the trial to permit a 1:1 computer generated randomization strategy for two groups receiving different treatment orders in a crossover design. An a priori power analysis revealed 46 participants would achieve > 90% power for a medium effect size of 0.5 for each outcome measure. Participants in the randomized, double-blind (including caretakers, children, experimenters and clincal assessors), placebo-controlled, crossover design were randomly allocated to receive intranasal OXT first followed by intranasal PLC or vice-versa. Additionally, study participants received a 2-week initial period of PLC intranasal sprays prior to their 6-week treatment period with OXT or continued PLC to allow assessment of possible expectation (placebo) effects [see 22]. The study was conducted in the Chengdu Maternal and Children’s Central hospital (CMCCH) between June 2019 and July 2021 in accordance with the guidelines for Good Clinical Practice and the principles of the Declaration of Helsinki and was reviewed and approved by the Ethics Committee of CMCCH (number 201983) as well as the general ethics committee of the University of Electronic Science and Technology of China (number 1420190601). Parents or legal guardians of all participants provided written informed consent and the trial was pre-registered (Chinese Clinical Trial Registry: ChiCTR1900023774: https://trialsearch.who.int/Trial2.aspx?TrialID=ChiCTR1900023774).

Eligible children were recruited from outpatient clinics at CMCCH and other children’s hospitals in Chengdu and Sichuan Province via specialist networks and in response to the trial registration. Participants were considered eligible if they were diagnosed with ASD according to the Diagnostic and Statistical Manual of Mental Disorders, Fifth Edition (DSM-V) [1]; aged 3–8 years and their diagnosis was confirmed with the Autism Diagnostic Observation Schedule - 2 (ADOS-2) [53] administered by two research reliable authors. No participants were receiving any psychotropic medications during the course of the trial. We excluded individuals with any identified genetic or chromosomal abnormalities (Fragile X or Rett syndromes) or diagnosed with a neurological disease (e.g., epilepsy, cerebral palsy), or psychiatric disorder other than ASD or severe respiratory, hearing or visual impairments. Out of 84 children screened, 54 were considered eligible but caregivers of 8 declined to participate leaving a total of 46 (see Fig. 1A for crossover trial CONSORT [60]).

**Fig. 1.**
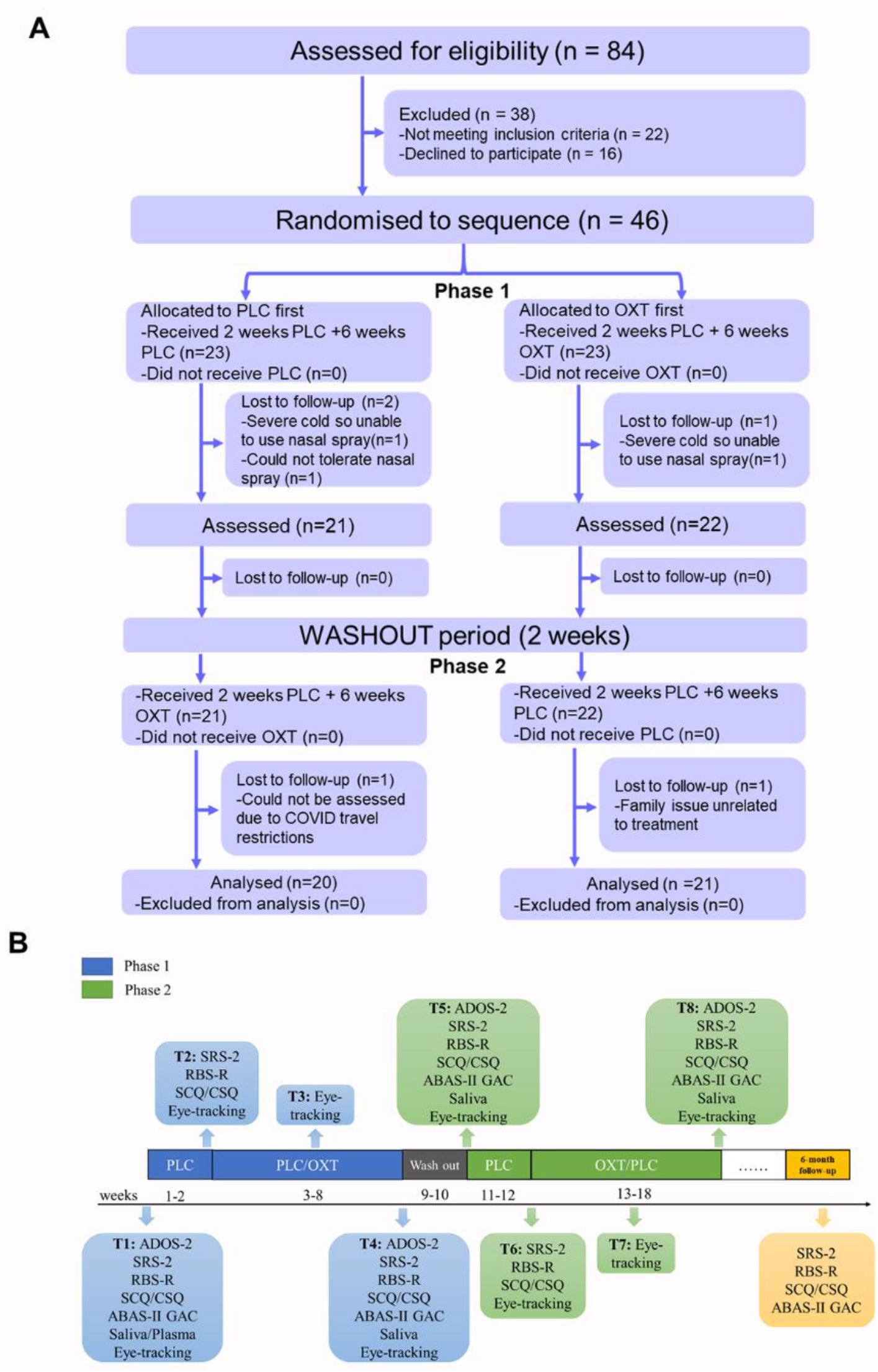
Trial Protocol. A. CONSORT flow diagram for crossover trials [60] and B. Schematic showing the organization of the two phases of the cross-over trial and intranasal treatments. Subjects received either oxytocin (OXT) or placebo (PLC) in each phase for 6 weeks and received PLC in the 2 weeks prior to this. There was a 2 week wash out period between phases. Outcome measures were taken at 8 different time-points during the trial (T1 – T8) and at a 6-month follow-up. Autism Diagnostic Observation Schedule – 2 (ADOS-2); Social Responsivity Scale-2 (SRS-2); Adaptive behavior assessment system-II (ABAS-II); Social communication quotient (SCQ); Repetitive behavior scale - revised (RBS-R); caregiver strain questionnaire (CSQ); Eye-tracking, Tasks 1 and 2. Points where blood or saliva samples were taken are also indicated.

### Assessment and medication schedules

For the trial protocol see Figure 1B. Following informed consent by a caregiver, eligible children visited CHCCM for recording pre-treatment outcome measures before their caregiver administered the first nasal spray under supervision. Caregivers were then instructed to administer the nasal spray in the early morning every other day during the treatment phases and to engage their child in a positive social interaction session for 30 minutes. Details of instructions and composition of OXT and PLC sprays and masking are provided in online supplementary information.

### Primary outcome measures

Primary outcome measures were the gold-standard clinical assessment using ADOS-2 scores (Comparison and Total scores) [53] and the SRS-2 [54,55] total score in line with previous trials [19,22,30]. The comparison ADOS-2 score was included since both modules 1 and 2 were used [53], although each individual was always assessed using the same ADOS-2 module. ADOS-2 and SRS-2 subscale scores were only anayzed if total scores showed significant improvements.

### Secondary outcome measures

Secondary outcome measures were chosen which had some incremental value to the main primary ones in line with recent recommendations [61]. These included a general assessment of adaptive behavior, (Adaptive behavior assessment schedule - ABAS-II) [62] and more detailed specfic assessments of social communication (Social communication quotient - SCQ) [63] and repetitive behaviors (Repetitive behavior scale- revised - RBS-R) [64]. These secondary outcome measures can distinguish ASD and typically developing children, are sensitive to autism symptom severity and have good internal consistency. As such, they adhere to general recommendations for using clinimetric criteria [61]. Children additionally completed two eye-tracking tasks which provide an objective assessment of social attention and can also discriminate between ASD and typically developing children [13,14,56-58] (see online supplementary information). Finally, the strain experienced by caregivers themselves was assessed using the Cargeiver strain questionnaire (CSQ) [65]. Blood (6 ml) or saliva samples [66] were taken at different time points for OXT measurement (see online supplementary information).

### Safety and other caretaker assessed behavioral observations

Caregivers recorded adverse events and any examples of positive or negative behavioral changes daily during treatment phases using an 18-item checklist (see supplementary Table S2) and could discuss any issues with medical staff or investigators during assessment visits (i.e. every 2-3 weeks).

### Autism sub-type and oxytocin receptor genotype

The social sub-type of children was assessed using the validated Beijing Autism Sub-type Questionnaire (BASQ - aloof, passive or active but odd sub-types) [see 59]. Buccal cell samples were taken for analysis of *OXTR* genotype [17], including SNPs rs53576, rs2254298, rs2268491, rs2268498, rs237887 based on their reported associations with autism [16,17]

### Statistical analysis

All statistical analyses were performed using SPSS version 22.0 (SPSS Inc., Chicago, IL, USA). An initial analysis of scores on questionnaires (SRS-2, SCQ, RBS-R, CSQ) completed both before and after the initial lead-in 2-week control PLC treatment revealed no significant differences (see online supplementary Table S1) and so were averaged to form a single baseline score. Given the within-subjects design, measures after each treatment phase were expressed as differences from baseline. The effectiveness of the wash-out period to prevent carry-over effects was also assessed (see also online supplementary information). A general linear model (GLM) approach was adopted for analysis of differences scores with main effects of treatment (OXT vs PLC) reported. Based on an initial analysis of ADOS-2 difference scores, the model incorporated age, gender and treatment order as covariates. To control for possible floor or ceiling effects in outcome measures, basal scores/measures were also included as covariates as well as basal plasma OXT concentrations due to a previous trial reporting these influenced treatment outcome [19]. Effect sizes were calculated using partial η^2^ (large >0.14, medium >0.06, small >0.01) [67]. Multiple comparisons within main primary and secondary outcome measures were controlled for using a False-Discovery Rate (FDR) Benjamini-Hochberg correction [68]. For the two eye-tracking paradigms repeated measures ANOVAs were performed. An additional more robust clinical assessment of improvement was performed on primary outcome measures by calculating a Reliable Change Index (RCI) [69] which indicates whether the observed change is significantly greater than a difference that could have occurred due to random measurement error alone. Differences in numbers of subjects identified with reliable improvement during OXT relative to PLC were analysed using McNemar’s chi-squared test. Correlations between outcome measures and altered OXT concentrations were assessed with Pearson tests. Missing data was only imputed (using average differences for other subjects) for saliva OXT concentrations under PLC where three subjects had one missing sample.

To assess the potential long-term impact of the intervention, caregivers completed some of the questionnaire-based measures (SRS-2, ABAS-II, SCQ, RBS-R and CSQ) at a 6-month follow up. Raw outcome measure scores were compared with those at the end of the OXT and PLC phases of the initial intervention using the GLM model.

Daily frequencies of urination and defecation were compared between treatments using Wilcoxon tests. Numbers of participants exhibiting non-serious adverse events in the OXT and PLC phases of the trial were compared statistically using chi-square tests for paired categorical data. In all cases p < 0.05 two-tailed was considered significant.

## Results

### Participants, baselines and assessment of carry-over effects

Five subjects did not complete both phases of the trial, leaving a total of 41 included in the final analysis (21 in the OXT-first arm of the trial and 20 in the PLC-first)(see Fig. 1A). Demographic details and baseline measures are provided in Table 1. The absence of significant treatment carry-over effects in the cross-over design was confirmed by demonstrating the absence of differences in baselines of outcome measures before each of the two treatment phases (see online supplementary Table S2 and Figure S1). Additionally, treatment order was included as a covariate in all analyses of treatment effects.

**Table 1.** Demographic and baseline measures Demographic, baseline clinical and questionnaire measures and basal oxytocin (OXT) concentrations in all 41 autistic children and those receiving OXT first followed by placebo or placebo first followed by OXT. Data for measures number or mean and standard deviation in brackets. Statistic and p-values show no differences in baseline measures between the sub-groups with different treatment orders. Autism Diagnostic Observation Schedule – 2 (ADOS-2); Social Responsivity Scale-2 (SRS-2); Adaptive behavior assessment system-II, global adaptive composite (ABAS-II GAC), Social communication quotient (SCQ), Repetitive behavior scale - revised (RBS-R), caregiver strain questionnaire (CSQ) and Beijing Autism Subtype questionnaire (BASQ). Statistic: t-tests, *chi-square (Fisher’s Exact Test) or ^#^ Mann-Whitney test.

| Variable | Total | Oxytocin first | Placebo first | Statistic | P |
| --- | --- | --- | --- | --- | --- |
| N | 41 | 21 | 20 |  |  |
| Age (years) | 5.0(1.3) | 5.1(1.4) | 4.9(1.2) | 0.618 | 0.540 |
| Sex: Male/female | 38/3 | 20/1 | 18/2 |  | 0.606* |
| **ADOS-2** |  |  |  |  |  |
| **Comparison score** | 6.9(1.0) | 6.7(1.1) | 7.1(1.0) | -1.291<sup>#</sup> | 0.197 |
| **Total score** | 16.9(4.1) | 16.6(4.7) | 17.3(3.5) | -0.557 | 0.581 |
| Social affect | 12.6(3.6) | 12.0(3.9) | 13.2(3.2) | -0.992 | 0.327 |
| Restricted and repetitive behavior | 4.3(1.7) | 4.5(1.8) | 4.2(1.6) | 0.700 | 0.488 |
| **SRS-2 total** | 97.7(22.2) | 99.8(22.7) | 95.6(21.9) | 0.614 | 0.543 |
| Social awareness | 13.5(3.3) | 13.8(3.1) | 13.2(3.5) | 0.567 | 0.574 |
| Social cognition | 18.8(4.0) | 19.1(4.2) | 18.4(3.8) | 0.535 | 0.596 |
| Social communication | 35.2(8.0) | 36.2(7.9) | 34.1(8.2) | 0.812 | 0.422 |
| Social motivation | 16.9(4.4) | 17.0(4.8) | 16.8(4.1) | 0.144 | 0.886 |
| Restricted interests and repetitive behavior | 13.4(6.1) | 13.8(6.1) | 13.1(6.2) | 0.410 | 0.684 |
| **ABAS-II GAC** | 63.90(13.56) | 62.43(11.11) | 65.45(15.88) | -0.709 | 0.483 |
| **SCQ** | 19.5(5.1) | 20.1(5.3) | 18.9(4.9) | 0.760 | 0.452 |
| **RBS-R** | 17.7(11.4) | 16.8(9.8) | 18.6(13.1) | -0.504 | 0.617 |
| **CSQ** | 8.3(2.4) | 8.3(1.8) | 8.3(2.9) | 0.052 | 0.959 |
| Base saliva OXT pg/ml | 8.4(5.1) | 7.8(4.0) | 9.2(6.2) | -0.793 | 0.433 |
| Base plasma OXT pg/ml | 6.4(3.9) | 7.0(4.9) | 5.8(2.3) | 0.992 | 0.327 |
| **Autism Subtype (BASQ)** |  |  |  |  |  |
| Aloof/ Passive /Active but odd | 18/18/5 | 7/11/3 | 11/7/2 |  |  |

### Analysis of primary outcome data

Table 2 and Figure 2A show that compared with PLC, OXT-treatment produced significant improvements in all primary outcome measures (ADOS-2 comparison score, pFDR = 0.006; ADOS-2 total score, pFDR < 0.001 and SRS-2 Total score, pFDR < 0.001 – all with large effect sizes, i.e. partial eta squared > 0.14). A secondary analysis of ADOS-2 subscales revealed improved social affect (pFDR = 0.021) but not restrictive and repetitive behavior (pFDR = 0.067), and for SRS-2 subscales for social awareness, cognition, communication and motivation (all pFDRs < 0.018) but not for restrictive and repetitive behavior (pFDR = 0.053).

**Table 2.** Primary clinical efficacy and secondary outcomes. Treatment outcome data from the 41 autistic children are mean and standard deviation (in brackets) difference scores for baseline compared to post-treatment (with either oxytocin or placebo) for primary outcome measures (in bold): Autism Diagnostic Observation Schedule – 2 (ADOS-2) – comparison score (Comp score) and total score and Social Responsivity Scale-2 (SRS-2) total score. Sub-scales for ADOS-2 (Social affect and restrictive and repetitive (Res and Rep) behavior and Social Responsivity Scale-2 (SRS-2): Social Aware, social awareness; Social Cog, social cognition; Social Comm, social communication; Social Motiv, social motivation and Res and Rep, restrictive interests and repetitive behavior are also given. Secondary outcome measure questionnaires: Adaptive behavior assessment system-II, global adaptive composite (ABAS-II GAC), Social communication quotient (SCQ), Repetitive behavior scale - revised (RBS-R) and caregiver strain questionnaire (CSQ) and % relative to baseline in saliva oxytocin (OXT) concentrations. Linear model (F-values) with age, gender, treatment order, plasma OXT and base scores as covariates and p values and effect sizes (partial η^2^) are also given. *** p<0.001, ** p<0.01 and *p<0.05 after false discovery rate (FDR) Benjamini-Hochberg correction for number of tests (i.e. 3 primary outcomes and 5 secondary outcomes) or subscales of significant primary outcomes (2 for ADOS-2 and 5 for SRS-2).

| Measure | Oxytocin | Placebo | F-value | pFDR | Effect size |
| --- | --- | --- | --- | --- | --- |
| **Primary** |  |  |  |  |  |
| **ADOS-2** |  |  |  |  |  |
| **Comp Score** | **0.34(0.48)** | **-0.02(0.61)** | **10.59** | **.003\*** | **0.232** |
| **Total** | **1.34(1.62)** | **0.05(1.45)** | **14.10** | **<.001\*\*\*** | **0.287** |
| Social affect | 0.90(1.48) | 0.05(1.43) | 6.41 | .032* | 0.155 |
| Res and rep behavior | 0.44(1.30) | 0.00(0.84) | 3.58 | .067 | 0.093 |
| **SRS-2** |  |  |  |  |  |
| **Total** | **11.32(15.14)** | **-1.16(10.54)** | **19.59** | **<.001\*\*\*** | **0.359** |
| Social Aware | 1.30(2.91) | -0.83(2.50) | 12.58 | .002** | 0.264 |
| Social Cog | 2.34(3.34) | -0.65(3.54) | 13.82 | <.001*** | 0.283 |
| Social Comm | 4.29(7.05) | 0.62(5.25) | 6.48 | .018* | 0.156 |
| Social Motiv | 2.10(2.92) | -0.37(3.00) | 13.21 | <.001*** | 0.274 |
| Res and Rep | 1.28(3.46) | 0.06(2.79) | 4.01 | .053 | 0.103 |
| **Secondary** |  |  |  |  |  |
| ABAS-II GAC | 4.34(8.41) | -0.68(5.58) | 8.19 | .018* | 0.190 |
| RBS-R | 2.44(6.03) | -0.55(5.31) | 5.93 | .033* | 0.145 |
| SCQ | 0.70(3.67) | 0.56(3.39) | 0.03 | .860 | 0.001 |
| CSQ | 0.32(0.97) | 0.14(0.89) | 0.50 | .607 | 0.014 |
| OXT %inc | 191.81(105.43) | 95.49(26.43) | 29.50 | <.001*** | 0.480 |

**Fig. 2.**
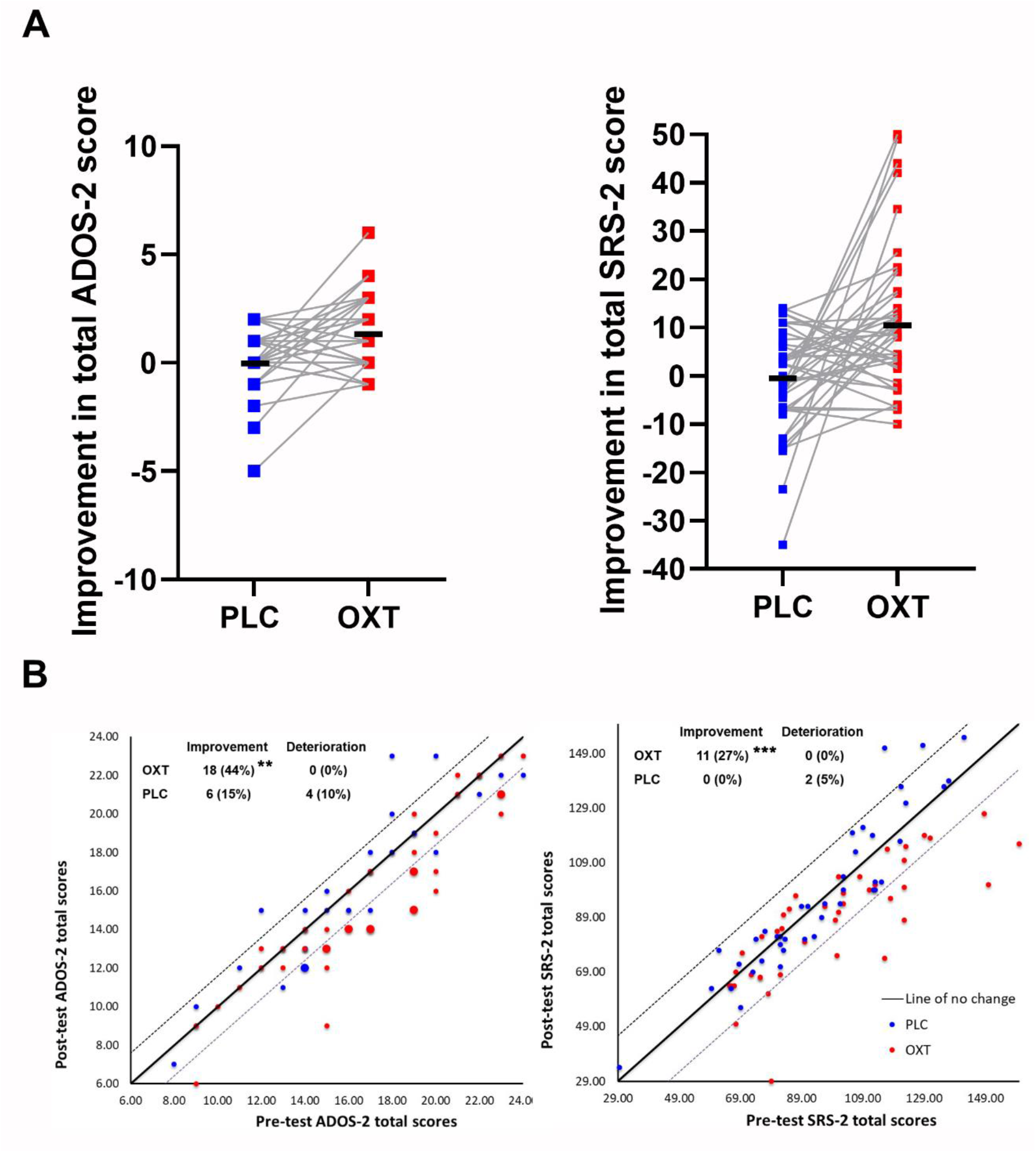
Changes in primary outcome measures. A. Plots showing differences between the PLC and OXT treatment effects in all 41 children for differences in Autism Diagnostic Observation Schedule – 2 (ADOS-2) and Social Responsivity Scale-2 (SRS-2) total scores. B. Reliable change index (RCI) plotted for both OXT and PLC treatment phases. The number of children showing reliable change (on or below lower dotted line) or deterioration (on or above upper dotted line) are given (also see online supplementary Table S3). Note: where two subjects have identical scores the size of the data dot is increased to indicate this. ** p < 0.01, *** p < 0.001 for OXT vs PLC McNemar’s chi-square.

An RCI analysis showed that 44% (ADOS-2 total) and 27% (SRS-2 total) of individuals exhibited reliable improvement under OXT and none showed deterioration. Under PLC, 15% (ADOS-2 total) and 0% (SRS-2 total) showed reliable improvement and 10% (ADOS-2 total) and 5% (SRS-2 total) showed deterioration. The number of subjects exhibiting reliable improvement under OXT relative to PLC was significantly greater for ADOS-2 (chi-square = 7.2, p = 0.007) and SRS-2 (chi-square = 11, p < 0.001) total scores (see Fig. 2B and online supplementary Table S3).

### Analysis of secondary outcomes

Table 2 shows treatment outcome results for secondary outcome questionnaires and baseline saliva OXT concentrations. The OXT treatment compared to PLC improved ABAS-II global adaptive composite (GAC) (pFDR = 0.018) and RBS-R scores (pFDR = 0.033), and increased basal saliva OXT concentrations (pFDR < 0.001).

### Eye-tracking assessments

ANOVA analysis with treatment and time-point as factors only revealed a main effect of treatment on proportion of time spent viewing dynamic social (dancing children) compared to geometric patterns in Task 1 (F1,35 = 5.06, p = 0.031, partial η^2^ = 0.126, n = 36) indicating significant effects of OXT at both 3- and 6-week timepoints. For the Task 2 face emotion paradigm separate repeated measures ANOVA for mid- and post-treatment time points (only 30 subjects had data at both time points) only revealed a significant treatment x face region x face emotion interaction (F(6,228) = 4.113, p = 0.001, partial η^2^ = 0.010; n = 39) at the 6-week time point. Post-hoc tests revealed OXT significantly increased time spent viewing the eye region of angry (pFDR = 0.002), happy (pFDR = 0.027) and neutral (pFDR = 0.032) expression faces but decreased it for fearful faces (pFDR = 0.016) (see Fig. 3). Including time-point as an additional factor revealed a treatment x face emotion x time point interaction for the eyes (F(3, 93) = 4.116, p = 0.018, partial η^2^ = 0.117, n = 30) with post-hoc tests indicating OXT increased time viewing the eyes of angry faces at 6 compared to 3 weeks (pFDR = 0.03). There were no treatment effects on the absolute amount of time spent viewing stimuli in either task (see online supplementary information).

**Fig. 3.**
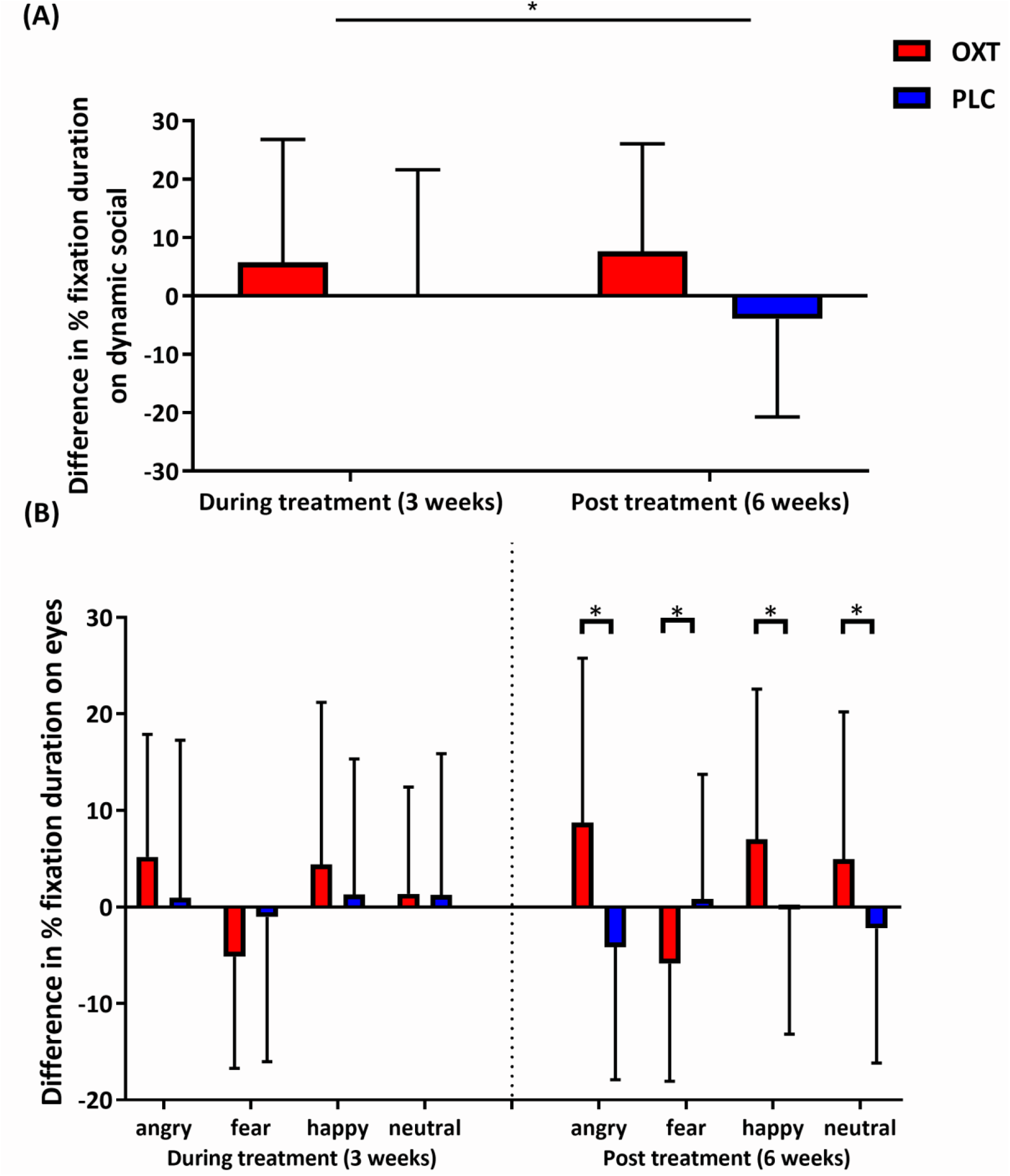
Treatment effects on two eye-tracking paradigms (A) Task 1 - mean ± SD treatment (oxytocin – OXT and placebo PLC) differences in % total duration viewing the dynamic social compared to the dynamic geometric stimuli and (B) Task 2 the eye region of all, angry, happy, fearful and neutral faces. Differences are shown for both during treatment (after 3-weeks) and post-treatment (after 6-weeks). **p<0.01. *p<0.05 post-hoc FDR corrected tests for OXT vs PLC.

### Parental assessments

Parental observations during treatment also supported symptom improvement during OXT relative to PLC (see online supplementary information).

### Potential factors influencing oxytocin-induced improvements

Increased saliva OXT concentrations after OXT treatment phase were correlated with the improvement in SRS-2 (r = 0.401, p = 0.0126) and ABAS-II GAC (r = 0.604, p < 0.001) scores but not ADOS-2 total scores (r = 0.201, p = 0.197). Increased viewing the eyes of fearful faces was also correlated with change in basal OXT concentrations (r = -0.327, p = 0.045, see online supplementary Figure S2).

Improvements in both primary and secondary outcome measures were similar across the different autism social sub-types (see online supplementary Tables S4 and S5) indicating that OXT improved symptoms independent of sub-type. For OXTR genotype only the rs2268491 SNP showed preliminary evidence for modulating effects of OXT with greater SRS-2 improvements and OXT concentration changes in CC-relative to T+-carriers, although this did not survive FDR correction (see online supplementary Table S6).

### Effects of intranasal oxytocin on safety and tolerability

No severe adverse effects were reported under either OXT or PLC treatments, although one participant withdrew during the PLC treatment phase due to being unable to tolerate the nasal spray (see Fig. 1A). For non-serious adverse events the only significant difference found was a small increase in the frequency of daily urination under OXT compared with PLC similar to a previous trial [22](see online supplementary Table S7).

### 6-month follow up

Caregivers of 27 children completed SRS-2, SCQ, RBS-R and CSQ and 30 completed the ABAS-II questionnaires remotely 6 months after treatment phases. None of the outcome scores at follow-up were worse than at the end of OXT treatment and the SRS-2 score remained significantly improved compared to the end of the PLC phase (see online supplementary Table S8). Thus, there was no evidence for the intervention having any long-term negative efffects.

## Discussion

Overall, the findings of this cross-over trial demonstrate that intranasal OXT treatment can significantly improve symptoms in young children with autism when given every-other-day as opposed to every day and followed by a period of positive social interaction. Importantly, the current trial not only achieved improvements in the same subjective measure (SRS-2) as reported in two previous studies in young children using daily treatment [19,22], but additionally for the first time found a significant reduction in the objective gold standard clinical measure, ADOS-2. Furthermore, the trial provided the first evidence for improved social functioning using two eye-tracking paradigms. Improvements were also confirmed from caregiver reports although they did not report reductions in their self-rated levels of strain (CSQ scores). At a 6-month follow up there was no indication that the intervention had any negative impact on symptom severity and reduced SRS-2 scores seen during OXT treatment were maintained. In agreement with previous trials [19,22,25,30], no serious adverse effects were found with the only significant change being an increase in urination frequency in line with a previous trial [22].

No previous PLC-controlled study in children has shown changes in total ADOS-2 scores in response to intranasal OXT [19,22] or to other pharmacological interventions such as the related peptide vasopressin [70] or bumetanide [71]. The improvement in ADOS-2 scores in the current trial had a large effect size and the more stringent RCI analysis indicated that a clinically reliable improvement was achieved in 44% of children and none showed deterioration. The 12-point reduction in SRS-2 total scores relative to PLC also had a large effect size with the RCI analysis indicating improvement in 27% of children. The number of children showing reliable improvement in both ADOS-2 and SRS-2 scores under OXT was also significantly greater than under PLC. Secondary analysis of ADOS-2 and SRS-2 subscales support the conclusion that OXT mainly improved social symptoms, most notably the ADOS-2 social affect subscale and the SRS-2, social awareness, cognition and motivation subscales. This was further supported by the two independent eye-tracking derived measures indicating that OXT increased visual attention to dynamic social relative to non-social stimuli and the eye-region of emotional faces. Only social communication showed inconsistent improvement under OXT with the SRS-2 communication subscale showing some improvement but not the SCQ secondary outcome measure. In terms of restrictive and repetitive behaviors both ADOS-2 and SRS-2 subscales showed a trend towards improvement under OXT although significance was only achieved in the more extensive RBS-R secondary outcome measure. Large-effect size improvements were also found in ABAS-II GAC scores which measure adaptive skills in conceptual and practical as well as social domains.

The eye-tracking secondary outcome measures demonstrated improved social attention following OXT treatment and may prove to be useful objective measures in future ASD clinical trials, particularly the dynamic social versus geometric pattern paradigm, or ‘GeoPref test,’ which is increasingly established as an early autism biomarker [56-58]. Interestingly, while OXT increased the proportion of time viewing the eye-region of happy, angry and neutral faces, it decreased that towards the eyes of fearful faces and so effects in children may be expression dependent. Increased time spent viewing the eye-region was only found after 6 weeks of treatment and not after 3 weeks. Thus, greater improvements may have occurred after a longer treatment period.

The two key innovations in the current trial were firstly to reduce the frequency of OXT intranasal doses to every-other-day and that caregivers engaged in positive social interaction/play with their children in the period immediately following intranasal administration. These innovations in combination may have contributed to greater improvements in autistic symptoms. At this point it is unclear whether the reduced dose-frequency or amount is of greatest importance. The average dose given every two days was between a quarter and a half of that in previous daily administration studies [19,22] and there is increasing evidence for inverted U-dose response curves for OXT in both preclinical [40,41] and clinical [72] studies. However, the pharmacodynamic advantage of giving a larger dose at less-frequent intervals is supported by the findings or our two preclinical studies [46-47]. A trial administering intranasal OXT to improve feeding and social skills in infants with Prader-Willi syndrome also reported significant effects using an every-other-day dosing schedule [73]. Mechanistically both a reduced dose or dose-frequency may have led to OXT primarily influencing its receptor’s excitatory Gq-coupled intracellular cascade more than its inhibitory Gi-coupled one [38-40]. Mechanisms of oppositional tolerance which can contribute to reduced efficacy of pharmacological treatments in depression might also be involved [74].

We did not observe evidence for improved symptoms when positive social interactions followed each PLC treatment and so it is unclear if there were additive or other types of interactions between the pharmacological and behavioral interventions, although we were careful to avoid differential placebo effects by having the same experimenter giving instructions to caregivers [see 75]. However, ensuring the children did not experience any negative social interactions immediately following OXT treatment may have been of most importance by avoiding its facilitation of their salience, leading potentially to social symptom impairment rather than improvement [see 52].

Our GLM analysis model included age, gender, treatment order, basal outcome measures and basal plasma OXT concentrations as covariates and none influenced observed main effects of treatment. Increases in saliva OXT concentrations were however significantly positively correlated with improvements in SRS-2 and ABAS-II GAC scores and negatively correlated with time spent looking at the eyes of fearful faces. Thus, individual differences in improved symptoms may have been influenced by the magnitude of increased basal OXT concentrations produced by the treatment. However, it should be noted that saliva OXT concentrations do not necessarily reflect those in cerebrospinal fluid [76] and individual differences in increased basal concentrations could be due to variable efficacy of nasal administration. There was preliminary evidence that OXT receptor genotype may influence treatment effects with the increase in basal OXTconcentrations and improvement in SRS-2 scores being less affected in T+ compared with CC carriers of rs2268491, although this did not pass FDR correction. A meta-analysis has identified rs2268491 as being associated with autism [16] and T+ carriers how a reduced neural response during an emotion recognition task [77]. Autism social sub-type (BASQ scores) did not influence improvements in primary or secondary outcome measures indicating that OXT had broadly similar effects across aloof, passive and active but odd sub-types [59].

A limitation of crossover designs is that they cannot reliably assess duration of effects. We did not find evidence for significant carry over effects between treatment phases but a future parallel design study could better assess the duration of OXT effects. Additionally, longer treatment durations may have achieved greater improvements in both social and repetitive behavior symptoms. Finally, the required period of positive social interaction/play between caretakers and children following intranasal treatments was not strictly controlled or structured in order to allow individual flexibility and did not itself result in significant improvements in symptoms. Future trials using more formal and structured positive behavioral interventions in combination with OXT treatment might result in greater symptom improvements.

## Conclusion

In summary, this cross-over trial on 41 young children with ASD has demonstrated that a 6-week intranasal OXT treatment using a novel infrequent dose regime followed by a period of positive social interaction is safe and improved core symptoms of ASD, particularly social functioning. The previous treatment philosophy of simply trying to increase basal OXT concentrations using more frequent large/escalating doses and without associating administration with positive social experience may have contributed to inconsistent findings in other clinical trials.

## Data Availability

All data for the Tables and Figures in the main paper will be provided in the final published version after peer review

## Acknowledgement

We thank the children and their families for their participation and support in this study.

## References

1. Diagnostic and Statistical Manual of Mental Disorders: DSM-V. (American Psychiatric Association, Washington, DC, 2013).

2. Lord C, Elsabbagh M, Baird G, Veenstra-Vanderweele J. Autism spectrum disorder. The Lancet. 2018; 392(10416):508–520.

3. Sun X, Allison C, Wei L, Matthews FE, Auyeung B, Wu YY, et al. Autism prevalence in China is comparable to western prevalence. Mol Autism. 2019; 10:7.

4. Gordon I, Brent C, Vander Wyk BC, Bennett RH, Cordeaux C, Lucas MV, et al. Oxytocin enhances brain function in children with autism. Proc. Natl. Acad. Sci. U. S. A. 2013; 110: 20953–20958.

5. Huang Y, Huang X, Ebstein RP, Yu R. Intranasal oxytocin in the treatment of autism spectrum disorders: a multilevel meta-analysis. Neurosci. Biobehav. Rev. 2021; 122:18–27.

6. Jiang X, Ma X, Geng Y, Zhao Z, Zhou F, Zhao W, et al. Intrinsic, dynamic and effective connectivity among large-scale brain networks modulated by oxytocin. Neuroimage. 2021; 227:117668.

7. Kendrick KM, Guastella AJ, Becker B. Overview of human oxytocin research. In Behavioral Pharmacology of Neuropeptides: Oxytocin. Curr Top in Behav Neurosci. 2017; 35:321–348.

8. Martins DA, Mazibuko N, Zelaya F, Vasilakopoulou S, Loveridge J, Oates A, et al. Effects of route of administration on oxytocin-induced changes in regional cerebral blood flow in humans. Nat. Commun. 2020; 11:1160.

9. Andari E, Duhamel JR, Zalla T, Herbrecht E, Leboyer M, Sirigu A. Promoting social behavior with oxytocin in high-functioning autism spectrum disorders. Proc Natl Acad Sci USA. 2010; 107:4389– 4394.

10. Auyeung B, Lombardo MV, Heinrichs M, Chakrabarti B, Sule A, Deakin JB. Oxytocin increases eye contact during a real-time, naturalistic social interaction in males with and without autism. Transl Psychiatry. 2015; 5(2):e507.

11. Le J, Kou J, Zhao W, Fu M, Zhang Y, Becker B, et al. Oxytocin biases eye-gaze to dynamic and static social images and the eyes of fearful faces: associations with trait autism. Transl Psychiatry. 2020; 10:1–12.

12. Le J, Zhao W, Kou J, Fu M, Zhang Y, Becker B, et al. Oxytocin facilitates socially directed attention. Psychophysiology. 2021; 58(9):e13852.

13. Black MN, Chen NTM, Iyer KK, Lipp OV, Bölte S, Falkmer M, et al. Mechanisms of facial emotion recognition in autism spectrum disorders: Insights from eye tracking and electroencephalography. Neurosci Biobehav Rev. 2017; 80:488–515.

14. Chita-Tegmark M. Social attention in ASD: a review and meta-analysis of eye-tracking studies. Res Dev Disabil. 2016; 48:79–93.

15. John S, Jaeggi AV. Oxytocin levels tend to be lower in autistic children: a meta-analysis of 31 studies. Autism. 2021; 25:2152–2161.

16. LoParo D, Waldman ID. The oxytocin receptor gene (OXTR) is associated with autism spectrum disorder. Mol Psychiatry. 2015; 20:640–646.

17. Montag C, Sindermann C, Melchers M, Jung, Luo R, Becker B, et al. A functional polymorphism of the OXTR gene is associated with autistic traits in Caucasian and Asian populations. Am J Med Genet B Neuropsychiatr Genet. 2017; 174:808–816.

18. Anagnostou E, Soorya L, Chaplin W, Bartz J, Halpern D, Wasserman S, et al. Intranasal oxytocin versus placebo in the treatment of adults with autism spectrum disorders: a randomized controlled trial. Mol Autism. 2012; 3:16.

19. Parker KJ, Oztan O, Libove RA, Sumiyoshi RD, Jackson LP, Karhson DS, et al. Intranasal oxytocin treatment for social deficits and biomarkers of response in children with autism. Proc Natl Acad Sci USA. 2017; 114:8119–8124.

20. Tachibana M, Kagitani-Shimono K, Mohri I, Yamamoto T, Sanefuji W, Nakamura A, et al. Long-term administration of intranasal oxytocin is a safe and promising therapy for early adolescent boys with autism spectrum disorder. J Child Adolesc Psychopharmacol. 2013; 23:123–127.

21. Watanabe T, Kuroda M, Kuwabara H, Aoki Y, Iwashiro N, Tatsunobu N, et al. Clinical and neural effects of six-week administration of oxytocin on core symptoms of autism. Brain. 2015; 138:3400–3412.

22. Yatawara CJ, Einfeld SL, Hickie IB, Davenport TA, Guastella AJ. The effect of oxytocin nasal spray on social interaction deficits observed in young children with autism: a randomized clinical crossover trial. Mol Psychiatry. 2016; 21:1225–1231.

23. Bernaerts S, Boets B, Bosmans G, Steyaert J, Alaerts K. Behavioral effects of multiple-dose oxytocin treatment in autism: a randomized, placebo-controlled trial with long-term follow-up. Mol Autism. 2020; 11:6.

24. Kosaka H, Okamoto Y, Munesue T, Yamasue H, Inohara K, Fujioka T, et al. Oxytocin efficacy is modulated by dosage and oxytocin receptor genotype in young adults with high-functioning autism: a 24-week randomized clinical trial. Transl Psychiatry. 2016; 6:e872.

25. Yamasue H, Okada T, Munesue T, Kuroda M, Fujioka T, Uno Y, et al. Effect of intranasal oxytocin on the core social symptoms of autism spectrum disorder: a randomized clinical trial. Mol Psychiatry. 2020; 25:1849–1858.

26. Dadds MR, MacDonald E, Cauchi A, Williams K, Levy F, Brennan J. Nasal oxytocin for social deficits in childhood autism: A randomized controlled trial. J Autism and Dev Disord. 2014; 44:521–531.

27. Guastella AJ, Gray KM, Rinehart NJ, Alvares GA, Tonge BJ, Hickie IB, et al. The effects of a course of intranasal oxytocin on social behaviors in youth diagnosed with autism spectrum disorders: A randomized controlled trial. J Child Psychol Psychiatry. 2015; 56(4):444–452.

28. Hirosawa T, Kikuchi M, Ouchi Y, Takahashi T, Yoshimura Y, Kosaka H, et al. A pilot study of serotonergic modulation after long-term administration of oxytocin in autism spectrum disorder. Autism Res. 2017; 10(5):821–828.

29. Munesue T, Nakamura H, Kikuchi M, Miura Y, Takeuchi N, Anme T, et al. Oxytocin for male subjects with autism spectrum disorder and comorbid intellectual disabilities: A randomized pilot study. Front Psychiatry. 2016; 7:2.

30. Sikich L, Kolevzon A, King BH, McDougle CJ, Sanders KB, Kim SJ, et al. Intranasal oxytocin in children and adolescents with Autism Spectrum Disorder. N Engl J Med. 2021; 385:1462–1473.

31. Rajagopal S, Shenoy SK. GPCR desensitization: acute and prolonged phases. Cell Signal. 2018; 41:9–16.

32. Gimpl G, Fahrenholz F. The oxytocin receptor system: structure, function, and regulation. Physiol Rev. 2001; 81:629–683.

33. Freeman SM, Ngo J, Singh B, Masnaghetti M, Bales KA, Blevins JE. Effects of chronic oxytocin administration and diet composition on oxytocin and vasopressin 1a receptor binding. Neuroscience. 2018; 392:241–251.

34. Pisansky MT, Hanson LR, Gottesman II, Gewitz JC. Oxytocin enhances observational fear in mice. Nat Commun. 2017; 8:2102.

35. Terenzi MG, Ingram CD. Oxytocin-induced excitation of neurons in the rat central and medial amygdaloid nuclei. Neuroscience. 2005; 134(1):345–354.

36. Leffa DD, Daumann F, Damiani AP, Afonso AC, Santos MA, Pedro TH, et al. DNA damage after chronic oxytocin administration is rats: a safety yellow light? Metab Brain Dis. 2017; 32:51–55.

37. Busnelli M, Chini B. Molecular basis of oxytocin receptor signalling in the brain: What we know and what we need to know. Curr Top Behav Neurosci. 2017; 35:3–29.

38. Chini B, Verhage M, Grinevich V. The action radius of oxytocin release in the mammalian CNS: From single vesicles to behavior. Trends Pharmacol Sci. 2017; 38:982–991.

39. Jurek B, Neumann ID. The oxytocin receptor: from intracellular signaling to behavior. Physiol Rev. 2018; 98:1805–1908.

40. Spengler FB, Schultz J, Scheele D, Essel M, Maier W, Heinrichs M, et al. Kinetics and dose dependency of intranasal oxytocin effects on amygdala reactivity. Biol Psychiatry. 2017; 82:885–894.

41. Martins DA, Mazibuko N, Zelaya F, Vasilakopoulou S, Loveridge J, Oates A, et al. Effects of route of administration on oxytocin-induced changes in regional cerebral blood flow in humans. Nat Commun. 2020; 11:1160.

42. Bales KL, Perkeybile AM, Conley OG, Lee MH, Guoynes CD, Downing GM, et al. Chronic intranasal oxytocin causes long-term impairments in partner preference formation in male Prairie voles. Biol Psychiatry 2013; 74(3):180–188.

43. Du P, He Z, Cai Z, Hao X, Dong N, Yuan W, et al. Chronic central oxytocin infusion impairs sociability in mandarin voles. Pharmacol Biochem Behav. 2017; 161:38–46.

44. Huang H, Michetti C, Busnelli M, Managò F, Sannino S, Scheggia D, et al. Chronic and acute intranasal oxytocin produce divergent social effects in mice. Neuropsychopharmacol. 2014; 39(5):1102–1114.

45. Winter J, Meyer M, Berger I, Royer M, Bianchi M, Kuffner K, et al. Chronic oxytocin-driven alternative splicing of Crfr2α induces anxiety. Mol Psychiatry. 2021; DOI: 10.1038/s41380-021-01141-x.

46. Kou J, Zhang Y, Zhou F, Sindermann C, Montag C, Becker, B. et al. A randomized trial shows dose-frequency and genotype may determine the therapeutic efficacy of intranasal oxytocin, Psychol. Med. 2020 Dec; 1–10 DOI: 10.1017/S0033291720003803.

47. Kou J, Zhang Y, Zhou F, Gao Z, Yao S, Zhao W, et al. Anxiolytic effects of chronic intranasal oxytocin on neural responses to threat are dose-frequency dependent. Psychother Psychosom. 2022; DOI: 10.1159/000521348.

48. Hurlemann R, Patin A, Onur OA, Cohen MX, Baumgartner T, Metzler S, et al. Oxytocin enhances amygdala-dependent, socially reinforced learning and emotional empathy in humans. J Neurosci. 2010; 30(14), 4999–5007.

49. Hu J, Qi S, Becker B, Luo L, Gao S, Gong Q, et al. Oxytocin selectively facilitates learning with social feedback and increases activity and functional connectivity in emotional memory and reward processing regions. Hum Brain Mapp. 2015; 36(6):2132–2146.

50. Zhuang Q, Zhu S, Yang X, Zhou X, Xu X, Chen Z, et al. Oxytocin-induced facilitation of learning in a probabilistic task is associated with reduced feedback- and error-related negativity potentials. J Psychopharmacol. 2021; 35(1):40–49.

51. Shamay-Tsoory SG, Abu-Akel A. The social salience hypothesis of oxytocin. Biol Psychiatry 2016; 79(3):194–202.

52. Ford CL, Young LJ. Refining oxytocin therapy for autism: context is key. Nat Rev Neurol 2022; 18: 67– 68.

53. Lord C, Rutter M, DiLavore PC, Risi S, Gotham K, Bishop S. Autism Diagnostic Observation Schedule – 2. 2nd edition. Los Angeles: Western Psychological Services; 2012

54. Constantino JN, Gruber CP. Social Responsiveness Scale – 2. 2nd Edition. Los Angeles: Western Psychological Services; 2012.

55. Shum KKM, Cho WK, Lam LMO, Laugeson EA, Wong WS, Law LSK. Learning how to make friends for Chinese adolescents with autism spectrum disorder: a randomized controlled trial of the Hong Kong Chinese version of the PEERS intervention. J Autism Dev Disord. 2019; 49:527–541.

56. Kou J, Le J, Fu M, Lan C, Chen Z, Li Q, et al. Comparison of three different eye-tracking tasks for distinguishing autistic from typically developing children and autistic symptom severity. Autism Res. 2019; 12:1529 – 1540.

57. Pierce K, Marinero S, Hazin R, McKenna B, Barnes CC, Malige A, et al. Eye tracking reveals abnormal visual preference for geometric images as an early biomarker of an autism spectrum disorder subtype associated with increased symptom severity. Biol Psychiatry. 2016; 79:657 – 666.

58. Wen TH, Cheng A, Andreason C, Zahiri J, Xiao Y, Xu R,et al. Large scale validation of an early-age eye-tracking biomarker of an autism spectrum disorder subtype. Sci Rep. 2022; 12:4253.

59. Meng FC, Xu XJ, Song TJ, Shou XJ, Wang XL, Han SP, et al. Development of an autism subtyping questionnaire based on social behaviors. Neurosci Bull. 2018; 34:789–800.

60. Dwan K, Li J, Altman D, Elbourne D. CONSORT 2010 statement: extension to randomized crossover trials. Br Med J. 2019; 366:14378.

61. Carrozzino D, Patierno C, Guidi J, Berrocal Montiel C, Cao J, Charlson ME, et al. Clinimetric criteria for patient-reported outcome measures. Psychother and Psychosom. 2021; 90:222–232.

62. Harrison P, Oakland T. Adaptive Behavior Assessment System, Second Edition (ABAS-II). 2nd Edition. Los Angeles. Western Psychological Services; 2008.

63. Rutter M, Bailey A, Lord C. The Social Communication Questionnaire: Manual. Los Angeles. Western Psychological Services; 2003.

64. Lam KS, Aman MG. The repetitive behavior scale-revised: independent validation in individuals with autism spectrum disorders. 2007; J Autism Dev Disord. 37:855–866.

65. Brannan A, Heflinger C, Bickman L. The caregiver strain questionnaire: measuring the impact on the family of living with a child with serious emotional disturbance. J Emotional Behav Disord. 1997; 5:212–222.

66. Fujii T, Schug J, Nishina K, Takahashi T, Okada H, Takagishi H. Relationship between salivary oxytocin levels and generosity in preschoolers. Sci Rep. 2016; 6:38662.

67. Cohen J, Cohen P, West SG, Aiken LS. Applied multiple regression/correlation analysis for the behavioral sciences. 2003; Third Edition. Routledge, New York.

68. Benjamini Y, Hochberg Y. Controlling the false discovery rate: a practical and powerful approach to multiple testing. J Roy Stats Soc B. 1995; 57 (1): 289–300.

69. Jacobson NS, Truax P. Clinical significance: a statistical approach to defining meaningful change in psychotherapy research. J Consult Clin Psychol. 1991; 59:12–19.

70. Parker KJ, Oztan O, Libove RA, Mohsin N, Karhson DS, Sumiyoshi RD. A randomized placebo-controlled pilot trial shows that intranasal vasopressin improves social deficits in children with autism. Sci Transl Med. 2019; 11:491.

71. Dai Y, Zhang L, Yu J, Zhou X, He H, Ji Y, et al. Improved symptoms following bumetanide treatment in children aged 3-6 years with autism spectrum disorder: a randomized, double-blind, placebo-controlled trial. Sci. Bull. 2020; 66:1591–1598.

72. Yamasue H, Kojima M, Kuwabara H, Kuroda M, Matsumoto K, Kanai C, et al. Effects of a novel nasal oxytocin spray with enhanced bioavailability on autism: a randomized trial. Brain. 2022; doi.org/10.1093/brain/awab291.

73. Tauber M, Boulanouar K, Diene G, Cabal-Berthoumieu S, Ehlinger V, Fichaux-Bourin MD, et al. The use of oxytocin to improve feeding and social skills in infants with Prader-Willi syndrome. Pediatrics. 2017; 139(2):e20162976.

74. Fava GA, Offidani E. The mechanisms of tolerance in antidepressant action. Prog Neuropsychopharmacol Biol Psychiatry. 2011; 35(7):1593–602.

75. Fava GA, Guidi J, Rafanelli C, Rickels K. The clinical inqadequacy of the placebo model and the development of an alternative conceptual framework. Psychother Psychosom 2017 Nov 3; 86:332–340.

76. Martin J, Kagerbauer S, Gempt J, Podtschaske A, Hapfelmeier A, Schneider G. Oxytocin levels in saliva correlate better than plasma levels with concentrations in the cerebrospinal fluid of patients in neurocritical care. J Neuroendocrinol. 2018; 30:e12596.

77. Uzefovsky F, Bethlehem RA, Shamay-Tsoory S, Ruigrok A, Holt R, Spencer M, et al. The oxytocin receptor gene predicts brain activity during an emotion recognition task. Mol Autism. 2019; 10:12.

