## Supplementary information for "Infrequent intranasal oxytocin followed by positive social interaction improves symptoms in autistic children: a pilot randomized clinical trial"

### Supplementary methods

### Additional details of trial procedures and protocols

**Nasal sprays, masking procedures and administration compliance measures:** Oxytocin (OXT) and placebo (PLC) nasal sprays were supplied sterile by a pharmaceutical company (Sichuan Meike Pharmaceutical Co, Sichuan, China) and contained the same components other than OXT (i.e. glycerol, sodium chloride and water). Intranasal spray bottles were labelled and distributed to caretakers by an individual not involved in any other aspect of the trial who was responsible for finally unmasking the treatment details at the end of the trial. Intranasal spay bottles for OXT or PLC were identical in appearance and labelling with each having a unique code. When caregivers and their children returned to CHCCM (every 2-3 weeks) in accordance with the protocol schedule (Fig. 1B), they were given new intranasal treatment bottles on each occasion and returned used ones for weighing as a measure of compliance. The volumes consumed in the intranasal; spray bottles did not differ significantly during the 6-week OXT (mean ± SD = 19.0 ± 3.6 ml) and PLC (18.9 ± 3.7 ml, t = 0.013, p = 0.897) treatment phases and were in line with the expected volume (21 doses during 6 weeks (i.e. 6 sprays each = 0.6 ml x 21 = 12.6 ml) + 1-3 priming sprays 0.1-0.3 ml each time; total volume = 14.7-18.9 ml).

**Post-nasal spray treatment social interaction protocol**

Caregivers of children participating in the trial were instructed to engage them in one to one positive social interactions for 30-60 minutes after nasal spray administration. They were instructed during these sessions to engage in mutual play with their child's favorite toy(s) or game and to encourage face to face and eye contact and mutual imitation (i.e. similar to the principles of the Early start Denver model [see 1]. All caregivers were given instructions on this by the same female experimenter to avoid possible confounds due to instructions being given by different individuals. While caregivers were not formally monitored for compliance they were informed that this activity in conjunction with nasal treatment was expected to have stronger beneficial effects.

**Eye-tracking measures and procedures**

Non-invasive eye-tracking (Tobii TX300, Tobii, Danderyd, Sweden) was performed at a 300 Hz sampling rate with a gaze accuracy of 0.4°. Recording and stimulus presentation and analysis were conducted using Tobii Pro Studio, E-prime 2.0 software, and E-Prime Extensions for Tobii (Psychology Software Tools, Pittsburgh, PA). The primary measure collected was total fixation duration with the proportion of time spent viewing the social stimulus in the first task paradigm (children dancing vs dynamic geometrical patterns) and the proportion of time spent viewing the eye, nose or mouth regions of the different emotional faces in the second paradigm. In both tasks trials were excluded from analysis where children spent < 5% of the time viewing stimuli as in [2] (Task 1, all trials included; Task 2, 1.6% of trials deleted).

During eye-tracking all children were either seated alone or on a caregiver's lap in front of a display screen on which all stimuli were presented. Initially, they were required to pass a five-point calibration using dynamic animations before being presented with the two experimental paradigms. Children were told to view the screen freely and caregivers instructed not to give any instructions to them about what to watch or interrupt them during the presentation period. Eye tracking measures were taken at all test time-points during the trial (see Fig. 1B). In the first Social Attention Task (Task 1) dynamic dancing Chinese children versus dynamic geometric patterns were displayed side by side similar to previous publications [2-4]. In the static emotional face task (Task 2) different individual emotional (angry, fear, happy and neutral) faces of either adults or children were presented. The whole eye tracking task lasted 114 seconds. Task 1 was similar to our previous study comparing autistic and typically children [2] and designed to measure differential attention to pairs of dynamic social (dancing individuals) and non-social (geometric patterns) images (in color; 545 × 430-pixel resolution). The dynamic social stimuli involved one, two or three children dancing, while the geometric patterns involved different moving shapes. The total presentation included 20 x 2s video clips prepared as two continuous videos. The stimuli (social vs. geometric) were presented simultaneously with one category on the left and the other on the right in one video and vice versa in the other, with the order being counter balanced across sessions. There was a 1500ms blank interval between the 2 videos. To reduce familiarity effects, four different sets of stimuli were presented once during each treatment phase although in a different order for each child. The static emotional face task (Task 2) contained 4 children's faces (boys vs. girls) and 2 adult faces (female vs. male) with angry, happy, fear and neutral expressions (in color and with a 680 × 845-pixel resolution and white background). Each face was presented for 2 seconds and followed by a 500ms interval where a black cross was displayed in the middle of a light grey background screen. In total, the face task lasted 72 seconds and the order of presented faces for each subject was random. As in Task 1, four different sets of faces were presented to subjects at different time points within each treatment phase. In total, there were 16 children's faces and 8 adult faces used (i.e. 6 faces for each of the 4 emotions).

**Blood and saliva samples and measurement of oxytocin**

Saliva samples were taken 24-48h after the end of treatment to avoid contamination by the final intranasal dose of OXT. Samples were collected onto ice and subsequently both plasma (after centrifugation) and saliva were stored at -80℃ until assayed for OXT within 6 months. Oxytocin concentrations were analyzed in duplicate with a widely used enzyme linked immunoassay (ELISA - ENZO Life Sciences, USA) [5,6]. A standard prior extraction step was performed in accordance with the manufacturers recommended protocol. Detection sensitivity was 3pg/ml, inter- and intra-assay variation was < 9%.

**Oxytocin receptor genotyping**

Following buccal cell sampling, oxytocin receptor genotyping for 5 SNPs (rs53576, rs2254298, rs2268491, rs2268498, rs237887) was performed using a Cobas Z 480 Light Cycler (Roche Diagnostics, Mannheim, Germany). A prior DNA extraction step was performed on a MagNA Pure 96 robot (Roche Diagnostics, Mannheim, Germany) and using probes designed by TIBMolBiol (Berlin, Germany) [see 7]

**Supplementary results**

**Analysis of potential carry over effects**

To assess the efficacy of the wash-out period between treatment phases we compared both the baselines at the beginning of each treatment phase as in a previous trial [8]. Table S2 shows that there were no significant differences between baselines for any primary or secondary outcome measures. Additionally, separate ANOVAs for raw ADOS-2 total and SRS-2 total scores for the two treatment orders (either OXT first or PLC first) and with time point as a factor revealed main effects of time point for ADOS-2 total scores for both sub-groups (OXT first- $F(3,60) = 4.941$, $p = 0.005$, partial $\eta^2 = 0.198$; PLC first – $F(3,57) = 5.652$, partial $\eta^2 = 0.229$) due to a significant post-hoc difference between pre- and post-OXT treatment time points (pFDR = 0.008 and 0.004 respectively). For SRS-2 there was a main effect of time for the PLC first subgroup ($F3,57 = 8.875$, $p < 0.001$, partial $\eta^2 = 0.318$) although only marginal for the OXT first subgroup ($F3,60 = 2.273$, $p =0.089$) with again the only significant post-hoc time points being pre- vs post- OXT (pFDR = 0.003 and 0.04 respectively) (see Figure S1). Finally, to further control for an influence of carry-over effects, treatment order was included as a nuisance covariate in the General Linear Model (GLM) used for all statistical analyses.

### Analysis of total viewing time of the stimuli in the two eye-tracking tasks

Task 1 mean ± SD = 31.03 ± 3.91s and 30.48 ± 5.66s out of 40s in PLC and OXT treatment phases respectively, p = 0.504 t-test; Task 2 mean ± SD = 5.71 ± 2.02s and 5.45 ± 2.77s out of 12s across the four face emotions in the PLC and OXT treatment phases respectively; p = 0.4416. There were no effects of OXT on total time viewing specific face emotions; all ps > 0.42).

### Caregiver feedback on positive and negative treatment effects on social behavior

Although anecdotal, and not a planned outcome measure, caregivers completed a weekly diary of observed positive and negative social behavior changes during the OXT and PLC treatment phases. Caregivers of 17/41 children reported incidences (1-6 occasions) of improved social behaviors during OXT treatment compared with 7/41 (1-2 occasions) during PLC (chi-square = 4.167, p = 0.041). Under OXT, caregivers mostly reported examples of improved social communication (n = 8) and playing with others (n = 6), although other examples included increased eye-contact/attention/smiling towards others, imitation, sharing and social hugging. In terms of examples of unusual negative anti-social behavior (other than irascibility and aggression reported in Table S7) only 3 caregivers reported this during PLC and 1 during OXT, each on a single occasion.

References:


1. Rogers SJ, Dawson G. Early start Denver Model for Children with Autism: 2020; Guildford Publishing

2. Kou J, Le J, Fu M, Lan C, Chen Z, Li Q, et al. Comparison of three different eye-tracking tasks for distinguishing autistic from typically developing children and autistic symptom severity. Autism Res. 2019; 12:1529 - 1540.
3. Pierce K, Marinero S, Hazin R, McKenna B, Barnes CC, Malige A, et al. Eye tracking reveals abnormal visual preference for geometric images as an early biomarker of an autism spectrum disorder subtype associated with increased symptom severity. Biol Psychiatry. 2016; 79:657 - 666.
4. Wen TH, Cheng A, Andreason C, Zahiri J, Xiao Y, Xu R, et al. Large scale validation of an early-age eye-tracking biomarker of an autism spectrum disorder subtype. Sci Rep. 2022; 12:4253.
5. Montag C, Sindermann C, Melchers M, Jung, Luo R, Becker B, et al. A functional polymorphism of the OXTR gene is associated with autistic traits in Caucasian and Asian populations. Am J Med Genet B Neuropsychiatr Genet. 2017; 174:808-816.
6. Kou J, Lan C, Zhang Y, Wang Q, Zhou F, Zhao Z, et al. In the nose or on the tongue? contrasting motivational effects of oral and intranasal oxytocin on arousal and reward during social processing. Trans. Psychiatry. 2021; 11: 94.
7. Fujii T, Schug J, Nishina K, Takahashi T, Okada H, Takagishi H. Relationship between salivary oxytocin levels and generosity in preschoolers. Sci Rep. 2016; 6:38662.
8. Yatawara CJ, Einfeld SL, Hickie IB, Davenport TA, Guastella AJ. The effect of oxytocin nasal spray on social interaction deficits observed in young children with autism: a randomized clinical crossover trial. Mol Psychiatry. 2016; 21:1225-1231.

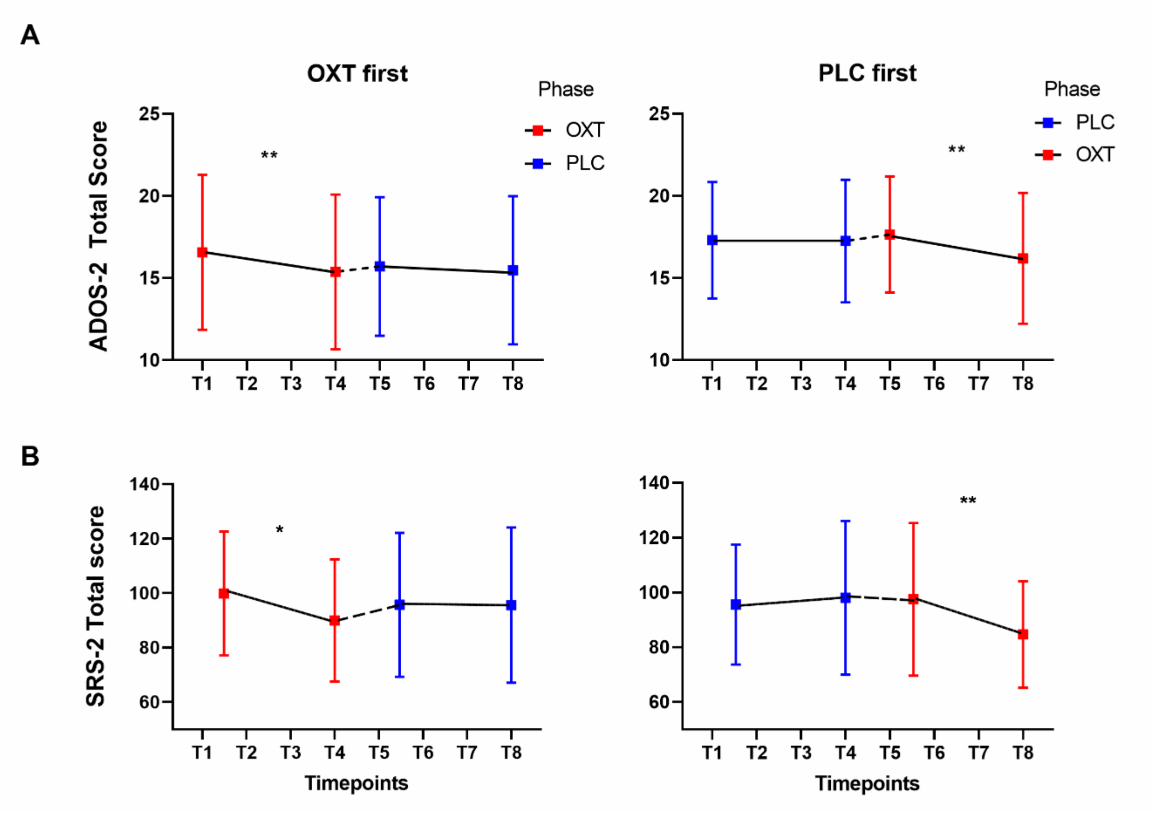


Fig. S1. Changes in primary outcome measures over time. Mean ± standard deviation scores for (A) Autism Diagnostic Observation Schedule – 2 (ADOS-2) total and (B) Social Responsivity Scale-2 (SRS-2) total at different time points during the trial for the subjects receiving oxytocin (OXT) first (n = 21) or placebo (PLC) first (n = 20). The SRS-2 was measured on two occasions at the beginning of each phase (T1 and T2 and T5 and T6) when all subjects were receiving PLC. No treatment occurred between time points 4 and 5 (wash out). * $p < 0.05$ or ** $p < 0.01$ post-hoc FDR corrected tests between 2 time points.

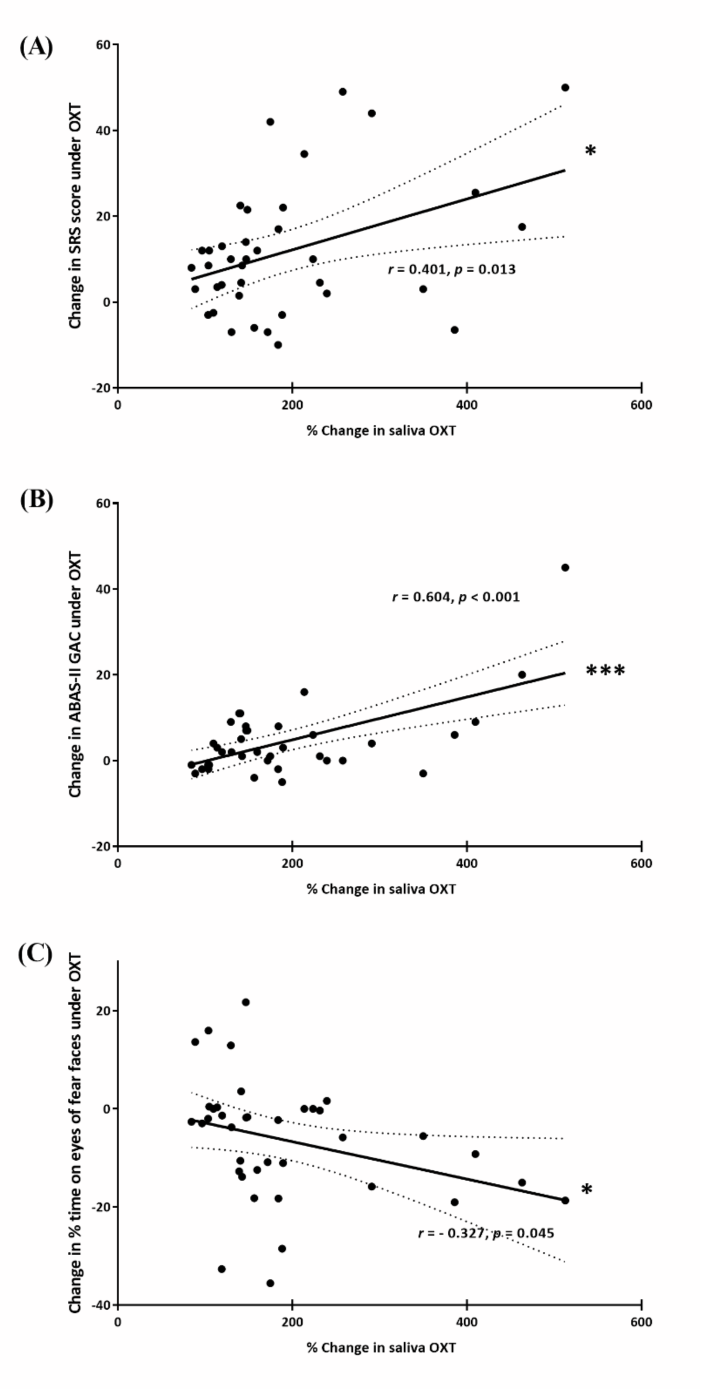


Fig. S2. Correlations between increased saliva oxytocin (OXT) concentrations and improved outcome measures. Regression graphs plot improvements in (A) Social responsivity scale (SRS-2) total scores and (B) Adaptive behavior assessment system-II, Global Adaptive Composite (ABAS-II GAC) scores positively associated with % change in basal saliva OXT concentrations at the end of the OXT treatment phase together with r and p-values (Pearson) (C) shows the same but for a negative association between time spent looking at the eyes of fearful faces and % change in basal OXT.

### Table S1 Baselines for outcome measures prior to treatment phases 1 and 2

| | Phase 1 | | | Phase 2 | | |
|---|---|---|---|---|---|---|
| **Measure** | Baseline 1 | Baseline 2 | t/p | Baseline 1 | Baseline 2 | t/p |
| **Primary** | | | | | | |
| SRS-2 Total | 97.85(20.78) | 97.63(25.48) | 0.10/0.921 | 96.39(25.88) | 96.73(28.51) | 0.236/0.815 |
| **Secondary** | | | | | | |
| RBS-R | 18.73(12.18) | 16.61(11.87) | 1.797/0.080 | 17.17(14.32) | 16.49(13.76) | 0.731/0.469 |
| SCQ | 19.78(5.04) | 19.22(5.59) | 1.211/0.233 | 18.39(5.64) | 18.39(6.31) | 0.00/1.00 |
| CSQ | 8.45(2.43) | 8.20(2.54) | 1.16/0.253 | 8.17(2.49) | 8.11(2.47) | 0.376/0.709 |

Baseline scores from the 41 autistic children who completed the study are mean and SD (in brackets) scores for the two baseline assessments taken prior to phase 1 and phase 2 treatments for primary and secondary measures: Social Responsivity Scale (SRS-2), Repetitive behavior scale - revised (RBS-R), Social communication quotient (SCQ) and caregiver strain questionnaire (CSQ). Test-statistic (t-values) and p values are given. There were no significant differences between the two baselines and so for the main analysis average values were used for the above measures.

**Table S2**
**Comparison of baseline measures before each treatment phase**

| Measure | Phase1 baseline | Phase2 baseline | F-value | pFDR |
|---|---|---|---|---|
| ADOS-2 Comp OXT first | 6.71(1.06) | 6.48(0.98) | 3.052 | 0.100 |
| ADOS-2 Comp PLC first | 7.10(0.97) | 7.20(1.01) | 1.185 | 0.294 |
| ADOS-2 Total OXT first | 16.57(4.72) | 15.71(4.22) | 5.101 | 0.076 |
| ADOS-2 Total PLC first | 17.30(3.54) | 17.65(3.53) | 1.209 | 0.289 |
| SRS-2 - OXT first | 99.83(22.74) | 95.69(26.43) | 0.611 | 0.446 |
| SRS-2 - PLC first | 95.55(21.90) | 97.48(27.90) | 0.493 | 0.493 |
| ABAS-II GAC - OXT first | 62.43(11.11) | 67.57(16.45) | 4.332 | 0.108 |
| ABAS-II GAC - PLC first | 65.45(15.88) | 66.55(17.95) | 0.644 | 0.435 |
| SCQ - OXT first | 20.10(5.33) | 18.60(5.83) | 2.077 | 0.169 |
| SCQ - PLC first | 18.88(4.93) | 18.18(5.54) | 0.894 | 0.359 |
| RBS-R - OXT first | 16.79(9.78) | 14.88(9.29 | 1.650 | 0.217 |
| RBS-R - PLC first | 18.60(13.11) | 18.88(17.24) | 0.037 | 0.850 |
| CSQ - OXT first | 8.35(1.76) | 8.26(1.93) | 0.115 | 0.739 |
| CSQ - PLC first | 8.31(2.92) | 8.01(2.96) | 1.596 | 0.226 |
| OXT pg/ml - OXT first | 8.13(2.91) | 7.84(4.05) | 0.190 | 0.668 |
| OXT pg/ml - PLC first | 9.16(6.18) | 7.43(4.98) | 1.468 | 0.247 |

Comparison of baselines for outcome measures before each treatment phase to assess potential carry-over effects. Data are mean SD for the two groups of participants receiving different treatment orders (Oxytocin (OXT) first followed by placebo (PLC) (n=21) or PLC first followed by OXT (n=20). Baselines for SRS-2, SCQ, RBS-R and CSQ are an average of 2 baseline scores taken before and after a 2-week PLC lead-in treatment. For ADOS-2 Comparison (Comp) and Total scores and ABAS-II GAC scores and saliva OXT concentrations the baseline is prior to the PLC lead-in treatment. Autism Diagnostic Observation Scale -2 (ADOS-2) Social Responsivity Scale (SRS-2), Adaptive behavior assessment system-II, global adaptive composite, ABAS-II GAC, Social communication quotient, SCQ, Repetitive behavior scale - revised (RBS-2) and caregiver strain questionnaire (CSQ). General Linear Model (F-values) and pFDR values are given. There were no significant differences between baselines.

**Table S3**

**Reliable change index (RCI) clinical analysis of outcome measures**

| Measure | Oxytocin n(%) | | | Placebo n(%) | | |
|---|---|---|---|---|---|---|
| | **Reliable deterioration** | **Uncertain change** | **Reliable improvement** | **Reliable deterioration** | **Uncertain change** | **Reliable improvement** |
| ADOS-2 | 0 (0%) | 23 (56%) | 18 (44%)** | 4 (10%) | 31 (75%) | 6 (15%) |
| SRS-2 | 0 (0%) | 30(73%) | 11 (27%)*** | 2 (5%) | 39 (95%) | 0 (0%) |

Reliable change index analysis of the numbers and percentage (in brackets) of participants in the trial identified as showing reliable improvement or deterioration or uncertain change after the oxytocin and placebo treatment phases. ADOS-2 = Autism Diagnostic Observation Schedule -Second Edition. Test re-test reliability = 0.98. SRS = Social Responsiveness Scale. Test re-test reliability = 0.94. ** $p < 0.01$ and *** $p < 0.001$ for oxytocin vs placebo McNemar's chi-square.

**Table S4 Autism subtype effects on primary and secondary outcomes**

| Measure | Aloof | Passive | Active but odd | F-value | P |
|---|---|---|---|---|---|
| N | 18 | 18 | 5 | | |
| **ADOS-2** | | | | | |
| Comparison score | 0.28(0.96) | 0.39(0.61) | 0.80(0.45) | 2.935 | 0.231 |
| Total score | 1.00(2.20) | 1.11(1.81) | 2.20 (3.19) | 0.624 | 0.541 |
| Social affect | 0.89(2.52) | 0.56(1.65) | 1.00(2.35) | 0.144 | 0.866 |
| Restricted and repetitive behavior | 0.11(1.32) | 0.56(1.65) | 1.20(1.10) | 1.190 | 0.315 |
| **SRS-2 total** | 16.03(19.28) | ·110.86(24.75) | 5.50(11.87) | 0.568 | 0.571 |
| Social awareness | 1.06(3.47) | 2.97(4.88) | 3.00(2.03) | 1.13 | 0.333 |
| Social cognition | 3.72(5.03) | 2.44(4.74) | 2.30(8.66) | 0.297 | 0.745 |
| Social communication | 6.61(9.37) | 2.56(10.11) | -2.90(8.42) | 2.131 | 0.133 |
| Social motivation | 2.67(3.99) | 2.06(5.16) | 3.20(3.21) | 0.160 | 0.853 |
| Restricted interests and repetitive behavior | 1.97(3.76) | 0.83(5.77) | -0.10(3.70) | 0.476 | 0.625 |
| **ABAS-II GAC** | 3.28(6.01) | 5.78(15.62) | 5.40(5.08) | 0.132 | 0.876 |
| **SCQ** | 1.11(4.06) | -1.00(5.29) | 0.70(3.51) | 0.990 | 0.381 |
| **RBS-R** | 2.97(6.50) | 3.92(11.21) | -0.30(4.58) | 0.450 | 0.641 |
| **CSQ** | 0.08(1.38) | 0.18(1.33) | 0.54(1.09) | 0.229 | 0.796 |
| **OXT % inc** | 113.58(111.47) | 93.15(115.44) | 41.55(9.97) | 0.718 | 0.495 |

Treatment outcome data from the 41 autistic children divided into autism social sub-types (aloof, passive and active but odd). Data are mean and SD (in brackets) difference scores for oxytocin (OXT) relative to placebo treatment at the end of the 6-week treatment periods for the primary (ADOS-2 and SRS-2 and their sub-scales) and secondary (ABAS-II GAC, SCQ, RBS-R, CSQ) outcome measures as well as % increase in saliva OXT concentrations. Test-statistic (F-values) and p values for ANOVA analysis. There are no significant differences between treatment effects in the different sub-types.

**Table S5 Autism subtype and treatment effects in eye-tracking paradigms**

| Measure | Aloof | Passive | Active but odd | F-value | P |
|---|---|---|---|---|---|
| **N** | 18 | 18 | 5 | | |
| **Social vs Geometric** | | | | | |
| %Social | 18.53(27.34) | 2.69(20.13) | 18.90(30.40) | | |
| **Emotional Face** | | | | | |
| **%Eyes** | | | | | |
| Angry | 21.94(28.93) | 9.92(15.16) | 1.28(14.08) | | |
| Fear | -5.54(13.35) | -11.77(13.52) | 8.47(8.97) | | |
| Happy | 11.83(16.14) | 7.01(17.56) | -2.10(30.07) | | |
| Neutral | 13.98(23.86) | 3.41(18.77) | -3.93(17.15) | | |
| **%Nose** | | | | | |
| Angry | -8.64(23.57) | -5.40(17.10) | 5.06(9.82) | | |
| Fear | 5.73(27.07) | -1.94(25.85) | 9.81(30.94) | | |
| Happy | 1.90(28.71) | -0.43(19.72) | 6.51(17.45) | | |
| Neutral | -0.42(36.20) | -3.96(27.67) | 6.10(8.85) | | |
| **%Mouth** | | | | | |
| Angry | -0.67(16.19) | 1.54(23.15) | 6.22(13.53) | | |
| Fear | 5.52(28.28) | -0.75(25.61) | -8.50(25.88) | | |
| Happy | 10.94(19.05) | 4.54(23.88) | 5.60(25.07) | | |
| Neutral | -2.26(15.50) | 3.12(15.50) | -8.00(21.48) | | |
| Emotion*subtype | | | | 0.547 | 0.771 |
| Region*subtype | | | | 0.804 | 0.526 |
| Emotion*region*subtype | | | | 1.16 | 0.31 |

Treatment outcome data from the 41 autistic children divided into autism sub-types (aloof, passive and active but odd). Data are mean and SD (in brackets) difference scores for oxytocin relative to placebo treatment effects (i.e. (oxytocin – baseline) – (placebo – baseline)) at the end of the 6-week treatment periods for % total fixation duration for dynamic social relative to dynamic geometric and for face emotions % differences in total fixation time on the eye, nose, mouth and all face regions of the different faces. Statistic (F-values) and p values for ANOVA analysis of relevant interactions. There were no significant differences across sub-types.

**Table S6**
**Effect of oxytocin receptor genotype on primary outcomes and oxytocin increase**

| SNP | Genotype | ADOS-2 Comp OXT-PLC | p | ADOS-2 Tot OXT-PLC | p | SRS Total OXT-PLC | p | %OXT Inc OXT-PLC | p |
|---|---|---|---|---|---|---|---|---|---|
| rs53576 | AA (n=18) | 0.33(0.20) | .710 | 0.94(0.46) | .292 | 14.14(4.84) | .441 | 69.73 (16.23) | .325 |
| | G+ (n=21) | 0.43(0.16) | | 1.67(0.48) | | 8.98(4.52) | | 99.80 (25.30) | |
| rs2254298 | A+ (n=23) | 0.39(0.20) | .950 | 1.22(0.44) | .688 | 6.52(3.10) | .111 | 72.29 (18.88) | .286 |
| | GG (n=16) | 0.38(0.13) | | 1.50(0.54) | | 18.31(6.40) | | 106.47(26.11) | |
| rs2268491 | CC (n=17) | 0.35(0.15) | .798 | 1.29(0.55) | .807 | 20.29(6.37) | .044* | 145.76(33.88) | .020* |
| | T+ (n=24) | 0.42(0.18) | | 1.13(0.43) | | 6.94(3.01) | | 64.04 (15.92) | |
| rs2268498 | C+ (n=17) | 0.35(0.19) | .798 | 1.18(0.57) | .963 | 14.44(5.69) | .622 | 108.78 (29.88) | .550 |
| | TT (n=24) | 0.42(0.16) | | 1.21(0.42) | | 11.08(4.01) | | 87.25 (21.39) | |
| rs237887 | A+ (n=29) | 0.34(0.13) | .564 | 1.41(0.36) | .317 | 9.69(3.64) | .192 | 78.21 (18.44) | .106 |
| | GG (n=12) | 0.50(0.26) | | 0.67(0.74) | | 19.21(6.91) | | 140.76(38.21) | |

Primary treatment outcome data for the 41 autistic children as a function of oxytocin receptor (OXTR) genotype. An exploratory analysis was conducted for 5 different SNPs (numbers of subjects with each genotype are given in brackets). Data are mean and SEM (in brackets) difference scores for effects of oxytocin minus effects of placebo treatment (i.e (oxytocin – baseline) – (placebo – baseline)) for the primary outcome measures: Autism Diagnostic Observation Schedule – 2 (ADOS-2) – comparison score (Comp score) and additionally Total score; Social Responsivity Scale-2 (SRS-2) score and % increase in saliva OXT concentrations. T-test p values are also given. *$p<0.05$ uncorrected for the number of SNPs. With FDR correction for the number of SNPs, for rs2268491 SRS-Total pFDR = 0.220, %OXT pFDR = 0.100.

**Table S7**

**Reported physical and behavioral symptoms during treatments**

| | OXT | PLC | P-values |
|---|---|---|---|
| **Non-serious** | | | |
| **Physical** | | | |
| Urination | 5.74 (1.21) | 5.36 (1.14) | .007** |
| Defecation | 0.91 (0.49) | 0.88 (0.42) | .377 |
| Constipation | 6 (14.6%) | 6 (14.6%) | 1.00 |
| Diarrhea | 2 ( 4.9%) | 5 (12.2%) | .257 |
| Erythema | 4 ( 9.8%) | 4 ( 9.8%) | 1.00 |
| Fatigue | 10 (24.4%) | 9 (22.0%) | .819 |
| Fever | 3 ( 7.3%) | 8 (19.5%) | .132 |
| Headache | 0 ( 0.0%) | 1 ( 2.4%) | - |
| Insomnia | 1 ( 2.4%) | 4 ( 9.8%) | .180 |
| Loss of appetite | 14 (34.1%) | 19 (46.3%) | .384 |
| Polypnea | 1 ( 2.4%) | 2 ( 4.9%) | .564 |
| Rhinitis | 15 (36.6%) | 18 (43.9%) | .602 |
| Sore throat | 5 (12.2%) | 7 (17.1%) | .564 |
| **Behavioral symptoms** | | | |
| Aggression | 8 (19.5%) | 10 (24.4%) | .637 |
| Irritability | 16 (39.0%) | 18 (43.9%) | .732 |
| **Serious adverse events** | None | None | |

Data from 41 autistic children for either daily mean and standard deviation of frequencies of urination and defecation during the 6 weeks of either oxytocin (OXT) or placebo (PLC) treatment or numbers of children exhibiting specific symptoms on one or more occasions at any time during the 6-week treatment periods with percentage given in brackets. Data were obtained from weekly caretaker diary records. The p-values given are either for Wilcoxon tests comparing the frequencies of specific symptoms or Chi-square for numbers of children experiencing specific symptoms during the treatment periods under OXT compared to PLC. **$p<0.01$ two-tailed. No serious adverse effects were recorded.

**Table S8 Outcome measures at 6-month follow-up**

| Measure | Oxytocin | | | Placebo | | |
|---|---|---|---|---|---|---|
| | End of treatment | 6-month follow up | F/pFDR | End of treatment | 6-month follow up | F/pFDR |
| SRS-2 total | 89.59(19.14) | 91.00(25.71) | 0.198/ 0.661 | 98.89(27.93) | 91.00(25.71) | 11.795/ 0.004** |
| ABAS-II GAC | 69.10(18.36) | 69.07(19.33) | 0.001/0.980 | 65.93(15.95) | 69.07(19.33) | 2.510/0.126 |
| SCQ | 18.44(6.09) | 16.63(6.86) | 3.602/ 0.071 | 17.85(6.60) | 16.63(6.86) | 1.970/ 0.174 |
| RBS-R | 16.63(15.31) | 19.85(18.49) | 3.789/ 0.064 | 18.30(15.36) | 19.85(18.49) | 1.471/ 0.238 |
| CSQ | 8.15(2.71) | 8.49(2.83) | 1.222/ 0.282 | 8.24(2.76) | 8.49(2.83) | 1.825/ 0.191 |

Questionnaire outcome data (means and SD) for autistic children (n = 27 or n = 30 for ABAS-II) at 6-month follow up compared to the end of the treatment phase (OXT/ PLC) of the trial for a primary measure: Social Responsivity Scale-2 (SRS-2) and for secondary measures: Adaptive behavior assessment system-II, global adaptive composite, ABAS-II GAC, Social communication quotient, SCQ, Repetitive behavior scale - revised (RBS-R) and caregiver strain questionnaire (CSQ). General Linear Model (F-values) and p values are given. ** p < 0.01. Scores at the follow-up were not significantly different from those during the OXT treatment phase. SRS-2 scores remained significantly reduced compared with PLC treatment during the trial. Overall, therefore we found no evidence for any longer-term negative effects of the intervention.
